# *SECISBP2* Deficiency Causes a Lethal Perinatal Cardiomyopathy

**DOI:** 10.64898/2026.07.22.26358709

**Authors:** Carlos C. Smith-Diaz, Natasha Henden, Natalie Stewart, Samantha Bryen, Mark Graham, Claire Lawley, Alexandra Butters, Adam T. Piers, Amy Baker, David A. Elliott, Ebony Richardson, Edward Formaini, Enzo R. Porrello, Helen Doyle, Igor E. Konstantinov, Ingrid King, Inuli Subasinghe, Kathleen Le Marquand, Laura Catto, Laura Yeates, Lisa Ewans, Rani Sachdev, Rocio Rius, Samantha Ross, Sui Yu, Winnie She, Johan Duflou, Cas Simons, Daniel G. MacArthur, Felicity Collins, Ulrich Schweizer, Amali Mallawaarachchi, James McNamara, Jodie Ingles

**Author notes:** These authors contributed equally to the study (Carlos C. Smith-Diaz, Natasha Henden). **Correspondence:** Carlos Smith-Diaz, Jodie Ingles.

## Abstract

Selenoproteins are a specialised group of proteins that incorporate selenium, an essential micronutrient, in the form of selenocysteine. The SECIS binding protein 2 (SBP2), encoded by *SECISBP2*, is a crucial component of the selenocysteine incorporation machinery. *SECISBP2* deficiency compromises selenoprotein synthesis, and its knockout causes embryonic lethality in mice. Selenium is critical to cardiac function, and nutritional deficiency causes Keshan disease, a progressive cardiomyopathy. Biallelic variants in *SECISBP2* cause pleiotropic phenotypes including abnormal thyroid hormone metabolism, neurodevelopmental disorders and aortic aneurysms. No reported phenotypes to date include cardiomyopathy. We report a consanguineous South Asian family with a history of perinatal deaths due to progressive cardiomyopathy and intractable arrhythmias with a rare homozygous loss-of-function splice site variant in *SECISBP2*.

The *SECISBP2* c.1303-2A>G variant was homozygous in four affected offspring and heterozygous in the parents. One child without cardiac disease did not carry this variant. RNA sequencing confirmed that almost all transcripts would undergo nonsense-mediated decay. Further, we observed a pronounced decrease in *GPX1* and *SELENOH* selenoprotein mRNA, as well as a large decrease in SELENOH, GPX1, GPX3 and GPX4 cardiac protein abundance in homozygotes, a molecular hallmark of *SECISBP2* deficiency. Notably, we observed a >12-fold decrease in cardiac GPX4, a key selenoprotein that suppresses lipid peroxidation and ferroptosis, suggesting a possible ferroptosis-mediated mechanism for heart failure. Our analysis suggests that c.1303-2A>G is likely the most damaging homozygous variant discovered to date. For the first time, we show that *SECISBP2* is essential for human life, with almost complete loss-of-function causing a lethal perinatal cardiomyopathy characterised by pronounced cardiac selenoprotein loss.

## INTRODUCTION

In humans, selenoproteins comprise a family of 25 proteins that contain the rare amino acid, selenocysteine^1^. Selenoproteins are involved in diverse biological processes. For example, the glutathione peroxidases protect against oxidative damage by reducing lipid hydroperoxides, whereas iodothyronine deiodinases are critical for thyroid hormone metabolism^2^. Many selenoprotein-encoding genes or biosynthesis factors are associated with Mendelian disorders^3^. The selenoproteins are critical for cardiac and skeletal muscle function. Dietary selenium deficiency underlies the pathogenesis of Keshan disease, a severe and progressive cardiomyopathy^4^, whereas loss-of-function variants in *SELENON* cause autosomal recessive *SELENON*-related myopathy^5–7^. A deficiency in selenoprotein P, the main serum selenium carrier protein, is associated with an increased risk of heart failure^8^. Further, dilated cardiomyopathy or skeletal myopathy has been widely reported in selenium deficient patients on parenteral nutrition^9–13^ and reversible cardiomyopathy has also been documented following selenium supplementation in a patient with Crohn’s disease^14^.

Selenium incorporation into selenoproteins differs from that of other trace elements. Instead of being bound or enzymatically attached to the protein, selenium is inserted into the growing polypeptide during translation as selenocysteine. Selenocysteine is encoded by an in-frame UGA codon. While a UGA codon typically signals translation termination, in selenoprotein mRNAs, it is interpreted to direct the co-translational incorporation of selenocysteine^2,3,15^. Selenocysteine incorporation is governed by the selenocysteine insertion machinery. This includes a stem-loop RNA structure situated in the 3’ untranslated region of eukaryotic selenoprotein mRNA, known as the selenocysteine insertion sequence (SECIS) element^3^. Another essential component is the SECIS binding protein 2 (SBP2), a trans-acting regulatory factor encoded by *SECISBP2*, which was identified based on its ability to bind SECIS elements^16^. SBP2 plays a multifaceted role in selenoprotein synthesis by stabilising selenoprotein mRNA and facilitating the co-translational insertion of selenocysteine^17,18^.

*SECISBP2* deficiency causes a variety of symptoms. The first clinical report described a phenotype of elevated T4 (thyroxine) to T3 (triiodothyronine) ratios due to defective iodothyronine deiodination^19^. Subsequent reports have expanded the phenotypic spectrum to include neurodevelopmental disabilities, hearing impairment, hypotonia, muscle weakness, azoospermia and aortic aneurysms^20–22^. While previously reported cases of *SECISBP2* deficiency in humans are compatible with life, with presentation often in childhood or adolescence, total knockout of *Secisbp2* in mice causes embryonic lethality^17,18^.

Here, we report a family with multiple offspring affected by a lethal perinatal cardiomyopathy, in which we detected a novel homozygous *SECISBP2* variant in affected individuals. We sought to investigate the impact of the variant and whether it could account for the severe cardiac phenotype.

## METHODS

### Family Recruitment

Following the death of their fourth child, a consanguineous family of South Asian ancestry was recruited to Elusive Hearts, an Australian study aiming to identify novel causes of monogenic cardiovascular disorders. Eligibility is restricted to families residing within Australia who have undergone inconclusive clinical genetic testing for suspected hereditary cardiovascular disease. The Elusive Hearts study was approved by the Royal Children’s Hospital Human Research Ethics Committee (97521), and participants provided written informed consent. Medical records were obtained, and a detailed family history was collected. Case report ethics approval was also granted by the Sydney Children’s Hospitals Network Human Research Ethics Committee (CCR2026/9). Pedigree codes denote each family member as follows: father (I-1), mother (I-2), child 1 (II-1), child 2 (II-2), child 3 (II-3), child 4 (II-4), and child 5 (II-5).

### Whole Genome Sequencing

DNA from the parents and five deceased offspring underwent whole-genome sequencing (WGS). Trio WGS was first performed through the Victorian Clinical Genetics Services (I-1, I-2 and II-3). Additional genomes were sequenced by the Garvan Sequencing Platform (II-1, II-2 and II-4) and the Australian Genome Research Facility (II-5). WGS data processing was performed using the Centre for Population Genomics (CPG) CaRDinal platform following the DRAGEN GATK best practices. Reads were aligned to the hg38 reference genome using Dragmap (v1.3.0). Cohort-wide joint calling of single nucleotide variants (SNVs) and small insertion/deletion (indel) variants was performed using GATK HaplotypeCaller (v4.2.6.1) with “--dragen-mode” enabled. Variants were annotated using VEP 110. Structural variant (SV) calling was performed using GATK-SV^23^. The resulting callset was loaded into CPG’s CaRDinal deployment of *seqr* for analysis^24^. Mitochondrial variants were called from genome sequencing data using the GATK mitochondrial variant calling workflow with MuTect2 in mitochondria mode^25^, and outputs were processed to generate MitoReports for downstream review. Sample sex and relatedness quality checks were performed using Somalier (v0.2.15)^26^.

Candidate variant analysis was restricted to rare variants, defined as those present at <0.0001 mean allele frequency (MAF) and in zero homozygotes in gnomAD v2.1.1 or v3.1.2. We performed a genome-wide inheritance-driven analysis to identify homozygous and compound heterozygous SNVs and SVs, which co-segregated in all affected offspring and had a high impact Ensembl Variant Effect Prediction (VEP). We also performed a phenotype-driven analysis to identify any variant(s) present in at least one allele in any affected offspring across 555 genes with cardiac disease associations from PanelApp Australia. Rare SVs and SNVs with an Ensembl-VEP of high/moderate impact, or with a SpliceAI maximum raw delta score >0.10 were reviewed.

### RNA Sequencing and Sample Collection

Skin cells from an antemortem skin biopsy collected from II-3, and mesenchymal stem cells from an umbilical cord sample (UC-MSCs) collected from II-4, were cultured at the Children’s Hospital Westmead, Sydney. Cells were grown in Ham’s F-10 Nutrient Mix, HEPES (Gibco), supplemented with 10% Fetal Bovine Serum, 1% Penicillin-Streptomycin (5,000 U/mL) and 1% L-Glutamine (200 mM). Passage 2 fibroblasts and passage 1 UC-MSCs were cryopreserved following negative testing for mycoplasma. Peripheral blood samples collected in PAXgene RNA tubes from each parent were stored at -80°C prior to extraction and sequencing.

RNA was extracted from blood and frozen cells by Garvan Molecular Genomics, using the Qiagen PAXgene Blood RNA and Qiagen RNeasy Mini kits, respectively. The integrity of RNA from parental blood samples was assessed using the Tapestation system (Agilent Technologies) with RNA Integrity Number equivalent (RINe) scores of 8.4 and 9.1. The RNA integrity of tissue and control blood samples was assessed with the Qsep Bio-Fragment Analyzer, with an RNA Quality Number of 6.43 and 6.41 for tissues, and 8.61 and 8.62 for blood controls. Library preparation was performed using the Illumina stranded total-RNA with Ribo-Zero kit. Libraries were sequenced on the Element Biosciences AVITI instrument using a Medium Output 300-cycle kit and paired-end 150-cycle protocol.

FASTQ files were trimmed with TrimGalore! using the default parameters (Phred score ≥20). Alignment against GRCh38 was performed on the Gadi HPC system (NCI, Australia) using STAR v2.7.11b; --twopassMode Basic, --outSAMtype BAM SortedByCoordinate, --outSAMstrandField intronMotif. Sashimi plots were generated using Gviz. For selenoprotein mRNA analysis, matched tissue controls (fibroblast n=11; UC-MSC n=18) were obtained from the Gene Expression Omnibus (GEO) database. Controls comprised paired-end, reverse strand-specific libraries from primary cells from different donors, encompassing transcriptional variability due to differences in culture conditions, library preparation (polyA vs rRNA depletion) and sequencing protocols. Control FASTQ files were processed in the same manner as patient transcriptomes. Raw counts were quantified using featureCounts (minMQS = 20), filtering was performed to remove genes with zero counts, and differential gene expression analysis was performed using DESeq2 (FDR <0.05). The expression of positive and negative marker genes for UC-MSC and dermal fibroblasts was assessed to validate the cellular identity of controls.

### FFPE Cardiac Proteomics

Myocardial formalin-fixed paraffin-embedded (FFPE) tissue from II-1 was obtained during autopsy. Control myocardial FFPE samples were obtained from 6 deceased babies born at term with no cardiac abnormalities. Protein extraction from FFPE tissue was performed using an adapted protocol from Achter et al^27^.

Each FFPE scroll was digested with 800 µL lysis buffer [100 mM 4-(2-hydroxyethyl)-1-piperazineethanesulfonic acid (HEPES) pH 8.5, 5% sodium dodecyl sulfate (SDS) and 10 mM dithiothreitol (DTT)]. The samples were sonicated for 20 s (Branson, Sonifer 150 W, micro tip) and incubated at 100 °C for 80 min followed by incubation at 60 °C for 2 h. Tubes were centrifuged at 18,000 g for 20 min at 23 °C and the middle aqueous phase was collected. A second 10 min centrifugation was used to further remove debris. For each quarter extract, 60 µL of a 1:1 mixture of Sera-Mag Carboxylate Hydrophylic and Hydrophobic beads (Cytiva) was added and acetonitrile was added to 75% v/v. Beads were mixed for 15 min. Tubes were placed on a magnet and the supernatant was discarded. Three washes were performed, two with 75% acetonitrile and two with 100% acetonitrile. For proteomics, beads were resuspended in 80 µL 100 mM triethylammonium bicarbonate (TEAB) containing 5% acetonitrile and incubated for 5 min at 37°C. The solutions were made up to 10 mM DTT and incubated for 30 min at 37 °C, then alkylated with 20 mM 2-chloroacetamide for 30 min at 23 °C in the dark. To each suspension was added LysC (1:50 Fujifilm Wako) for digestion at 37 °C for 4 h, followed by digestion with trypsin (1:25, TrypZean, MERCK). The suspensions were placed on a magnet, the supernatant was collected, dried, resuspended in 10 µL TEAB and two additional digestions with trypsin were performed (1:25 at 37 °C for 6 h and 1:50 at 40 °C for 3 h). The solutions were acidified and prepared for LC-MS/MS injection using a STAGEtip^28^.

LC-MS/MS analysis was performed using a Vanquish Neo UHPLC system and Astral Orbitrap mass spectrometer (Thermo Fisher Scientific). The samples were loaded onto a Pepmap Neo C18 5 µm particle 5 mm long by 300 µm inside diameter trap column (Thermo Fisher Scientific) at up to 10 µL/min and at a maximum pressure of 800 bar. The sample was eluted through a 15 cm long 75 µm inside diameter Pepmap C18 EASY-spray column with 3 μm 100 Å particles (Thermo Fisher Scientific). The column was heated to 40 °C using the EASY-spray ion source operating a 1.9 kV. The S lens radio frequency level was 40 and capillary temperature was 280 °C.

The liquid chromatography used buffer A (solution of 0.1% formic acid) and buffer B (0.1% formic acid and 99.9% acetonitrile). After loading the sample in 2% buffer B, the gradient at 0.5 µL/min was to 6% buffer B in 0.5 min, to 22% buffer B in 34 min, to 31% buffer B in 2 min, to 90% buffer B in 0.6 min, held for 2 min and returned to 2 % B for 0.8 min. MS acquisition was for 40 minutes. The MS scans in the orbitrap analyser were at a resolution of 240,000 with automatic gain control set to 5,000,000 counts and a maximum ion time of 3 ms for m/z 380 to 980. The data-independent acquisition MS/MS scans with a window of 2 m/z in the Astral analyser had automatic gain control at 50,000 for a maximum ion time of 3 ms. The loop was controlled to 0.6 seconds. The MS/MS scan range was 150-2000 m/z. The normalised collision energy was 25.

The raw LC-MS/MS data was processed with DIA-NN v2.6. The *Homo sapiens* reference proteome (using one gene per protein) downloaded on June 12, 2026 from UniProtKB, with 20,652 genes, was used to create an *in silico* library, with contaminants included. Match between runs was enabled to activate the generation of an experimental spectral library, created from all samples. N-terminal methionine excision was allowed. Carbamidomethyl (C) was a fixed modification. Peptide length was 7-30. Precursor charge range was 1-4. Precursor and product ion m/z range matched the MS settings. Initial mass accuracy was 10 ppm and MS1 accuracy was 4.5 ppm. Digestion was set to trypsin/P with a maximum of 1 missed cleavage. Protein inference was enabled, as were all other default algorithm settings in DIA-NN v2.6. Around 4-5,000 proteins were identified depending on the sample, with comparable median protein intensities for all samples analysed.

Quantified protein abundances were filtered to remove proteins with zero abundance in any sample, yielding 4015 proteins, followed by log2 conversion and median centring. Data visualisation was performed in R.

### Literature Review

MasterMind Professional was used to identify published biallelic pathogenic variants in *SECISBP2*. Based on published functional studies, *in silico* splicing algorithms (SpliceAI, Pangolin), and natural splicing data (SpliceVault), variants were categorised as hypomorphic, with potential rescue from nonsense-mediated decay (NMD), or null (if no NMD rescue mechanism was identified).

## RESULTS

### Clinical and Family History

We report a consanguineous couple (first-degree cousins once removed) of South Asian ancestry with a longstanding history of pregnancy loss, including at least two miscarriages and a further five pregnancies that resulted in fetal, neonatal or infant death. The maternal medical history included polyendocrine metabolic ovarian syndrome (PMOS) and treated hypothyroidism. There was no notable paternal medical history. Both parents had unremarkable electrocardiograms (ECG) and transthoracic echocardiograms (TTE). Across the five pregnancies detailed below, the mother and father were 32-39 and 41-49 years of age, respectively.

#### Pregnancy 1 (II-1)

The male fetus was conceived naturally, with unremarkable first trimester screening. This pregnancy was complicated by gestational diabetes and group B *streptococcus* colonisation. The mother presented at 35+1 weeks’ gestation with concern of decreased fetal movements. Screening revealed a structurally normal fetal heart with arrhythmias and frequent ectopic beats. Ultrasonography two days later revealed fetal death *in utero*. Following induced labour, an autopsy was performed, which revealed bilateral pleural effusions indicative of early cardiac failure, and neuronal necrosis consistent with an acute cerebrovascular ischaemic event. Significant cardiac abnormalities were observed, including a dilated right ventricle with interstitial fibrosis, increased pulmonary and tricuspid valve ring circumferences, myocyte necrosis with hypereosinophilia and dystrophic calcification (**Figure 1**). The cause of death was attributed to complications of parvoviral myocarditis following positive PCR results.

**Figure 1:**
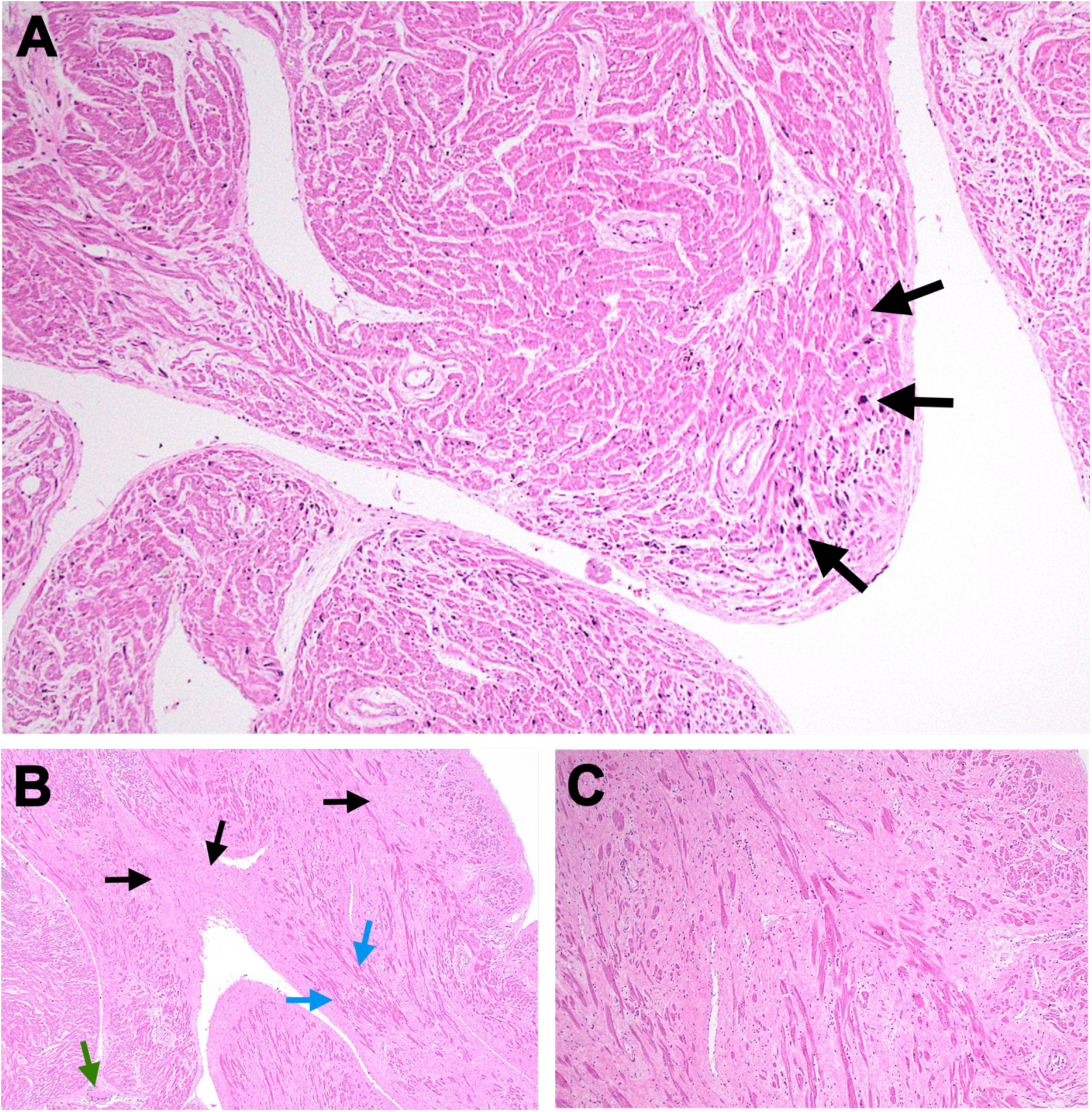
Cardiac Histology from the Autopsy of II-1. **A:** Hematoxylin & eosin stain, 100x original magnification. Section of endocardial region of ventricular myocardium showing subendocardial myocyte necrosis with hypereosinophilia and dystrophic calcification (arrows). **B:** Hematoxylin & eosin stain, 40x original magnification. Section of endocardial region of ventricular myocardium showing markedly diminished myocytes (blue arrows) with extensive areas of established fibrosis (black arrows). An area of subendocardial myocyte necrosis with hypereosinophilia and dystrophic calcification is depicted (green arrow). **C:** Hematoxylin & eosin stain, 100x original magnification. Section of left ventricular myocardium showing extensive replacement and interstitial fibrosis largely replacing myocardium.

#### Pregnancy 2 (II-2)

The female child was conceived naturally and presented *in utero* with hydrops fetalis and suspected intestinal atresia. Echocardiography revealed normal biventricular systolic function with a small mid muscular ventricular septal defect (VSD) with bidirectional shunt and no signs of cardiomyopathy. She subsequently developed severe combined immunodeficiency syndrome (SCID) and early onset inflammatory bowel disease and passed away in infancy. Clinical genetic testing revealed a novel pathogenic homozygous canonical splice site variant in the gene *TTC7A* (c.2356-1G>A), consistent with her phenotypic presentation of SCID and intestinal atresia.

#### Pregnancy 3 (II-3)

The third pregnancy, a male child, was conceived via *in vitro* fertilisation (IVF) with pre-implantation genetic testing (PGT) to exclude the pathogenic homozygous *TTC7A* variants identified in the previous pregnancy (II-2). Serial antenatal growth scans revealed cardiomegaly, a VSD and tricuspid regurgitation. The mother went into pre-term labour at 32-weeks’ gestation and delivery occurred via caesarean under general anaesthesia. The neonate was hypoxic and required intubation and ventilation. A chest x-ray taken shortly after birth showed cardiomegaly and likely hepatomegaly (**Figure 2A**). His echocardiogram was consistent with restrictive cardiomyopathy with marked bi-atrial enlargement and a moderate mid muscular VSD with a bidirectional shunt (**Figure 2B**). ECGs revealed interventricular conduction delay with repolarisation abnormalities, resting ST segment depression, inferior t-wave inversion and ventricular arrhythmias (**Figure 2C-D**). Other notable features included atypical facial features, and haematological abnormalities such as anaemia, thrombocytopaenia, and a blueberry muffin rash. A bone marrow biopsy was not diagnostic of a primary haematological disorder, and no cytogenetic abnormalities were observed. The neonate developed bradycardia and died shortly after birth. Clinical trio WGS was performed, identifying a maternally inherited heterozygous *TTN* c.50353A>T, p.(Arg16785Ter) variant. While considered a likely pathogenic variant, it was not considered the cause of the severe perinatal cardiomyopathy. No autopsy was performed.

**Figure 2:**
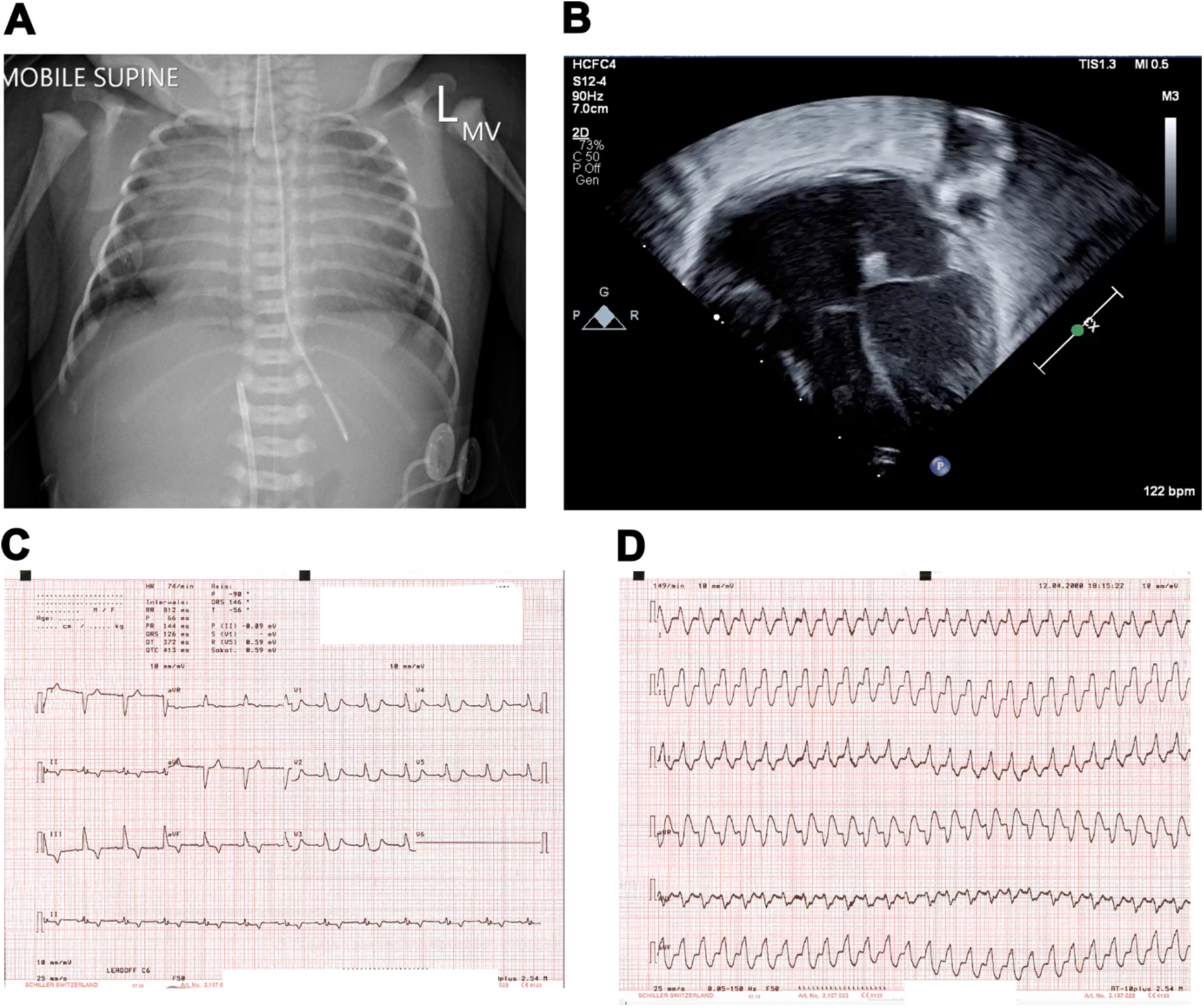
Radiographs, echocardiograms and electrocardiograms from II-3. **A:** Neonatal chest radiograph (anterior-posterior) demonstrating significant cardiomegaly and likely hepatomegaly. **B:** Transthoracic echocardiogram, apical four chamber view, demonstrating marked bi-atrial enlargement in the setting of restrictive cardiomyopathy. **C:** 12-lead ECG demonstrating a regular atrially led rhythm with marked interventricular conduction delay and abnormal repolarisation, resting ST segment depression and inferior t-wave inversion. **D:** 6-lead ECG demonstrating a broad complex arrhythmia.

#### Pregnancy 4 (II-4)

The fourth pregnancy, a female child, was conceived via IVF, with PGT performed to exclude the *TTC7A* and *TTN* variants. This pregnancy was complicated by gestational diabetes and obstetric cholestasis. Fetal echocardiography highlighted multiple VSDs as well as severe systolic dysfunction. The female neonate was born at 36 weeks’ gestation with a birth weight of 2300g, following caesarean under epidural anaesthesia. A chest x-ray demonstrated cardiomegaly and hepatomegaly (**Figure 3A**). ECGs revealed an abnormal axis, AV dissociation, interventricular conduction delay and abnormal repolarisation with resting ST segment abnormalities (**Figure 3B**). Echocardiography demonstrated persistent impaired left ventricular systolic function, bradycardia and multiple VSDs with bidirectional shunt (**Figure 3C**). Despite resuscitation she continued to deteriorate and passed from cardiogenic shock shortly after birth. No autopsy was performed.

**Figure 3:**
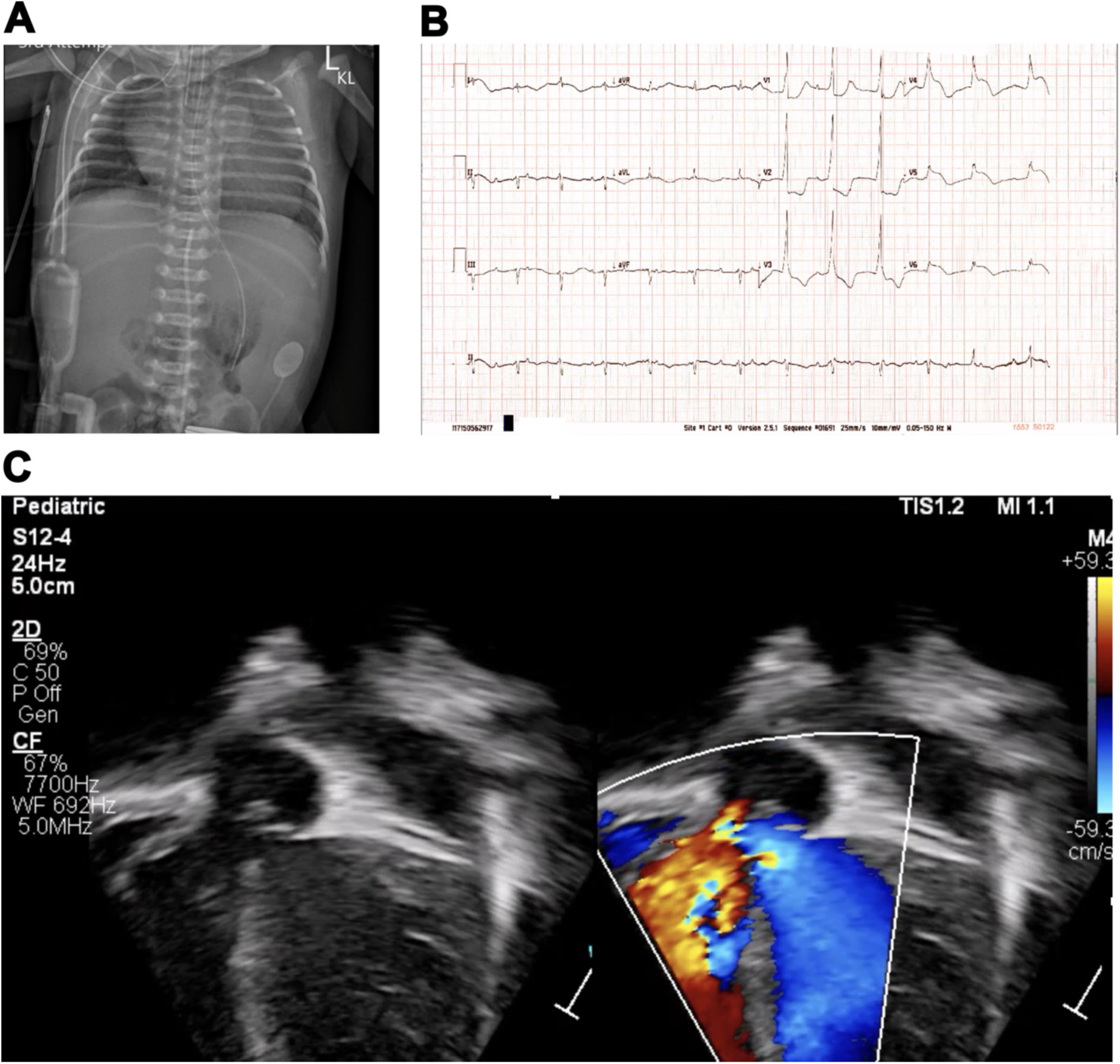
Radiographs, echocardiograms and electrocardiograms from II-4. **A:** Neonatal chest radiograph demonstrating cardiomegaly and hepatomegaly. **B:** 12-lead ECG demonstrating an abnormal axis, irregular rhythm with AV dissociation, interventricular conduction delay and abnormal repolarisation with resting ST segment abnormality. **C:** Still frame from transthoracic echocardiogram, apical four-chamber view, demonstrating severely impaired left ventricular systolic function, multiple VSDs with bidirectional shunt.

#### Pregnancy 5 (II-5)

The fifth pregnancy was conceived naturally and was closely monitored due to previous complications. By 19+5 weeks, a small muscular VSD was apparent. By 24+4 weeks, there was a moderate pericardial effusion with mild tricuspid regurgitation. At 28+4 weeks, cardiomegaly was identified, with mild tricuspid and mitral regurgitation and small pericardial and pleural effusions. At 31 weeks, severe hydrops with cardiomyopathy and atrial dilatation were identified. The fetus died *in utero* with worsening cardiac function and hydrops fetalis at 32+4 weeks after presenting with reduced fetal movements in the preceding days. No autopsy was performed.

### Genetic Analysis

We analysed WGS data from both parents and all five deceased children. **Figure 4** describes the family and genetic findings. A splice site variant, NM_024077.5(SECISBP2):c.1303-2A>G was homozygous in all offspring with the lethal cardiac phenotype (II-1, II-3, II-4, II-5), was absent from II-2 and was heterozygous in the parents. The heterozygous *TTN* truncating variant, NM_001267550.2(TTN):c.50353A>T,p.(Arg16785Ter), was inherited from the unaffected mother and was present in three of the four children with the lethal cardiomyopathy. This variant was reported as likely pathogenic, being a nonsense variant located in the asymmetric exon 267 (A-band) of the *TTN* meta-transcript, which is 100% spliced into cardiac titin isoforms, and absent from gnomAD v4, and ClinVar at the time of reporting. However, there is no evidence for heterozygous *TTN* truncations causing perinatal cardiomyopathies, and the variant did not segregate to all affected children. This variant was not, therefore, deemed the primary contributor to the cardiac phenotype. The *TTC7A* variant, NM_020458.4*(TTC7A)*:c.2356-1G>A, was homozygous in II-2, and heterozygous in I-1, I-2 and II-1.

**Figure 4:**
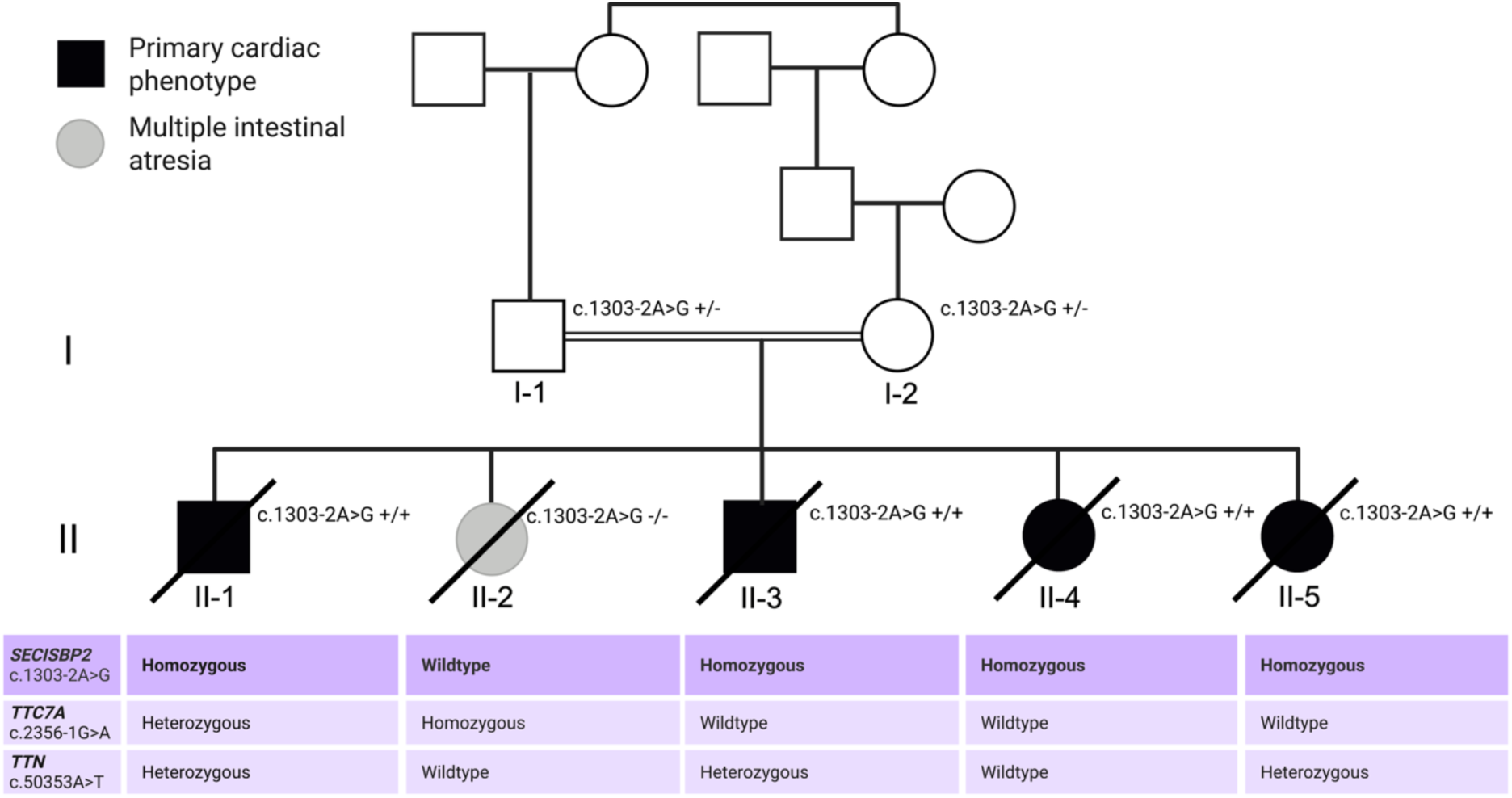
Pedigree and Genetic Variants. The *SECISBP2* c.1303-2A>G variant was homozygous in four offspring with cardiac disease (II-1, II-3, II-4, II-5) and was heterozygous in the parents (I-1, I-2). A fifth infant without cardiac disease did not carry the *SECISBP2* variant but instead had *TTC7A*-autosomal recessive combined immunodeficiency with multiple intestinal atresia and died in infancy (II-2).

### Variant Analysis

The *SECISBP2* c.1303-2A>G variant is located at the canonical splice acceptor site of exon 10 and is predicted to abolish normal splicing, resulting in a frameshift. The variant is present in two heterozygotes (at a frequency of 1.24 x 10^-6^) in the gnomAD v4.1 population database and has not been reported in the literature or ClinVar. SpliceAI suggested possible transcript rescue through an in-frame cryptic splice site within exon 10 at c.1326 (acceptor gain Δ0.18). We therefore used RNA sequencing to assess any potential rescue mechanisms.

Transcriptomes from two homozygous children (II-3 and II-4) showed a total loss of split reads between the splice donor and acceptor sites of exons 9-10, indicating a disruption of the exon 10 canonical splice acceptor site and exon 10 skipping (**Figure 5A and 5B**). We also observed the aberrant double skipping of exons 9-10 (**Figure 5A and 5B**). Only one split read was observed in the transcriptome of II-4 in support of a possible in-frame cryptic splice site (**Figure 5B**) with no supporting reads in the fibroblast transcriptome (**Figure 5A**). These data provide compelling evidence for out-of-frame exon 10 skipping, and very limited evidence for in-frame splicing rescue. We predict that skipping of the asymmetric exon 10 would cause a frameshift and a premature termination codon (PTC), leading to transcript degradation by NMD.

**Figure 5:**
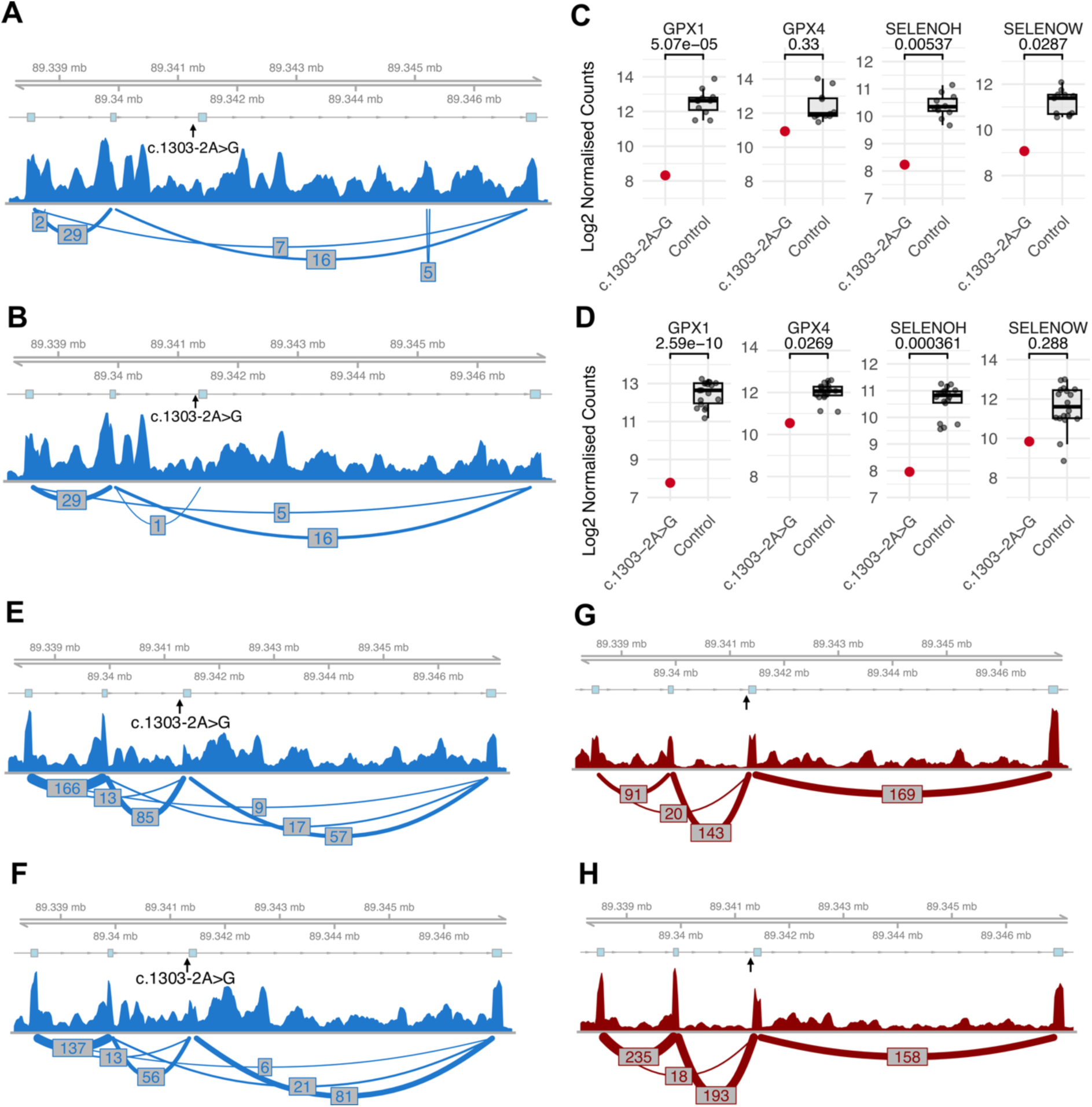
RNAseq Analysis. **A:** RNAseq data from fibroblasts from II-3 (c.1303-2A>G, homozygous) showing exon 10 skipping. **B:** RNAseq data from UC-MSCs from II-4 (c.1303-2A>G, homozygous) showing exon 10 skipping with one split read supporting a cryptic splice site. **C** and **D**: Log2 normalised counts and false detection rates from DESeq2 for *GPX1*, *GPX4*, *SELENOH* and *SELENOW* in II-3 fibroblasts (**C**) and II-4 UC-MSCs (**D**). **E and F:** Blood parental transcriptomes (c.1303-2A>G, heterozygous) (**E**) I-2, (**F**) I-1, showing an intermediate phenotype encompassing exon 10 skipping and normal exon 10 splicing. **G and H:** Blood transcriptomes from two *SECISBP2* wild-type individuals (confirmed with WGS). The minimum threshold for depicting a splice junction in blood transcriptomes was set at five split reads. No minimum threshold was set for fibroblasts or UC-MSCs.

We next compared the heterozygous transcriptomes with wild-type controls. Controls depicted normal exon 10 splicing (**Figure 5G and 5H**). In contrast, the heterozygous transcriptomes exhibited an intermediate splicing pattern with normal exon 10 splicing, exon 10 skipping and exon 9-10 double skipping (**Figure 5E and 5F**). Allelic imbalance in the heterozygous transcriptomes was indicative of intron retention, which would cause a frameshift, introduce a PTC and trigger NMD-degradation of affected transcripts. These predicted alternative gene products are summarised in **Table 1**. Interestingly, total *SECISBP2* mRNA expression was not decreased in homozygous patient cells when compared to matched tissue rRNA-depleted libraries from GEO. However, RNAseq expression data is not directly proportional to protein expression. We predict a dramatic decrease in SBP2 protein expression in homozygotes, due to the extremely high proportion of NMD-compliant transcripts that will be degraded by NMD or other mechanisms.

**Table 1:**
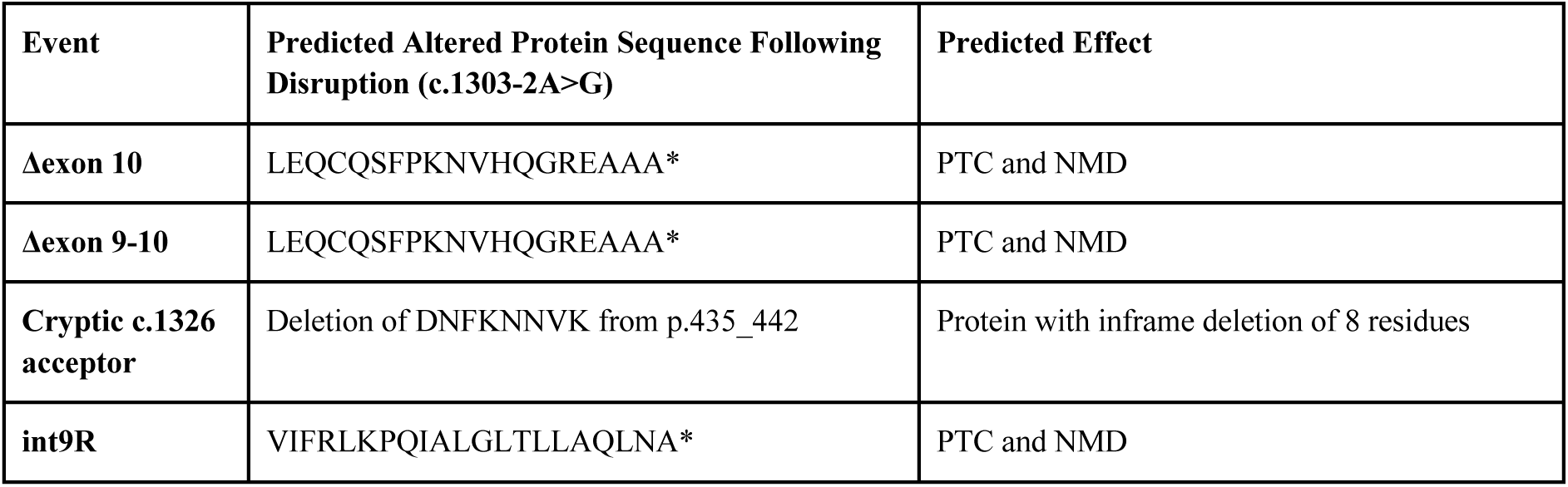
Predicted Gene Products (*SECISBP2* c.1303-2A>G)

### The Selenoprotein Transcriptome of Patient Cells

One of SBP2’s biochemical roles is stabilising selenoprotein mRNA and preventing mRNA degradation by NMD following UGA redefinition failure. However, not all selenoprotein mRNAs are equally susceptible to degradation following SBP2 loss. A core group of selenoproteins, notably *Gpx1*, *Selenow,* and *Selenoh*, are consistently downregulated in different mouse models of *Secisbp2* loss-of-function^17,18,29^. We therefore reasoned that these specific selenoprotein mRNAs would be decreased in patient cells carrying the homozygous c.1303-2A>G variant. Consistent with this theory, *GPX1* and *SELENOH* displayed a pronounced downregulation in cells from both homozygous offspring carrying the c.1303-2A>G variant (**Figure 5C and 5D**). The downregulation of *GPX1* was particularly robust with an FDR ≤ 5.07 x 10^-5^ and a log2 fold change > |-4| (> 16-fold downregulation) in cells from II-3 and II-4. This is consistent with *Gpx1* being the most affected selenoprotein mRNA in mouse *Secisbp2* knockout models^17,18^. Further, cells from one of the homozygous offspring displayed a downregulation of *SELENOW.* While the selenium status of controls is unknown, the large magnitude of decrease in *GPX1* and *SELENOH* mRNA, and relatively low control intersample variation, makes this comparison robust. Collectively, the downregulation of *GPX1, SELENOH* and *SELENOW* closely resembles the transcriptional changes observed in mouse models of *Secisbp2* loss-of-function^17,18,29^.

### Cardiac Proteomic Analysis

To assess the consequences of *SECISBP2* loss-of-function on cardiac selenoprotein abundance, we obtained FFPE myocardial tissue from II-1 and performed proteomic analysis using LC-MS/MS, incorporating tissue samples from six deceased term babies without cardiac disease as controls. Strikingly, the cardiac proteome of II-1 was characterised by a pronounced decrease in selenoprotein expression. The top downregulated protein overall was SELENOH, with a log2FC of -5.95 (>60x linear decrease). Other highly downregulated selenoproteins included GPX1 (Log2FC -5.06), GPX3 (Log2FC -4.49) and GPX4 (Log2FC -3.62) (**Figure 6A and 6B**). For these four selenoproteins, controls displayed low intersample variability with very large magnitude effect sizes. Other selenoproteins such as SELENOW and SEPHS2 were detectable in controls but totally absent in the proteome of II-1, suggesting either complete biological absence in II-1 or detection failure. SBP2 itself was not detectable in any sample with LC-MS/MS proteomics. Notably, HMOX1 (Heme oxygenase 1, Log2FC 2.92), GSTA1 (Glutathione S-transferase A1, Log2FC 2.35), MBOAT2 (Membrane-bound glycerophospholipid O-acyltransferase 2, Log2FC 4.57) and SQOR (Sulfide:quinone oxidoreductase, mitochondrial, Log2FC 1.75), proteins involved in oxidative stress responses, were also upregulated in II-1. We also observed an upregulation of FTH1 (Ferritin heavy chain, Log2FC 2.17) and FTL (Ferritin light chain, Log2FC 2.09) (**Figure 6C**).

**Figure 6:**
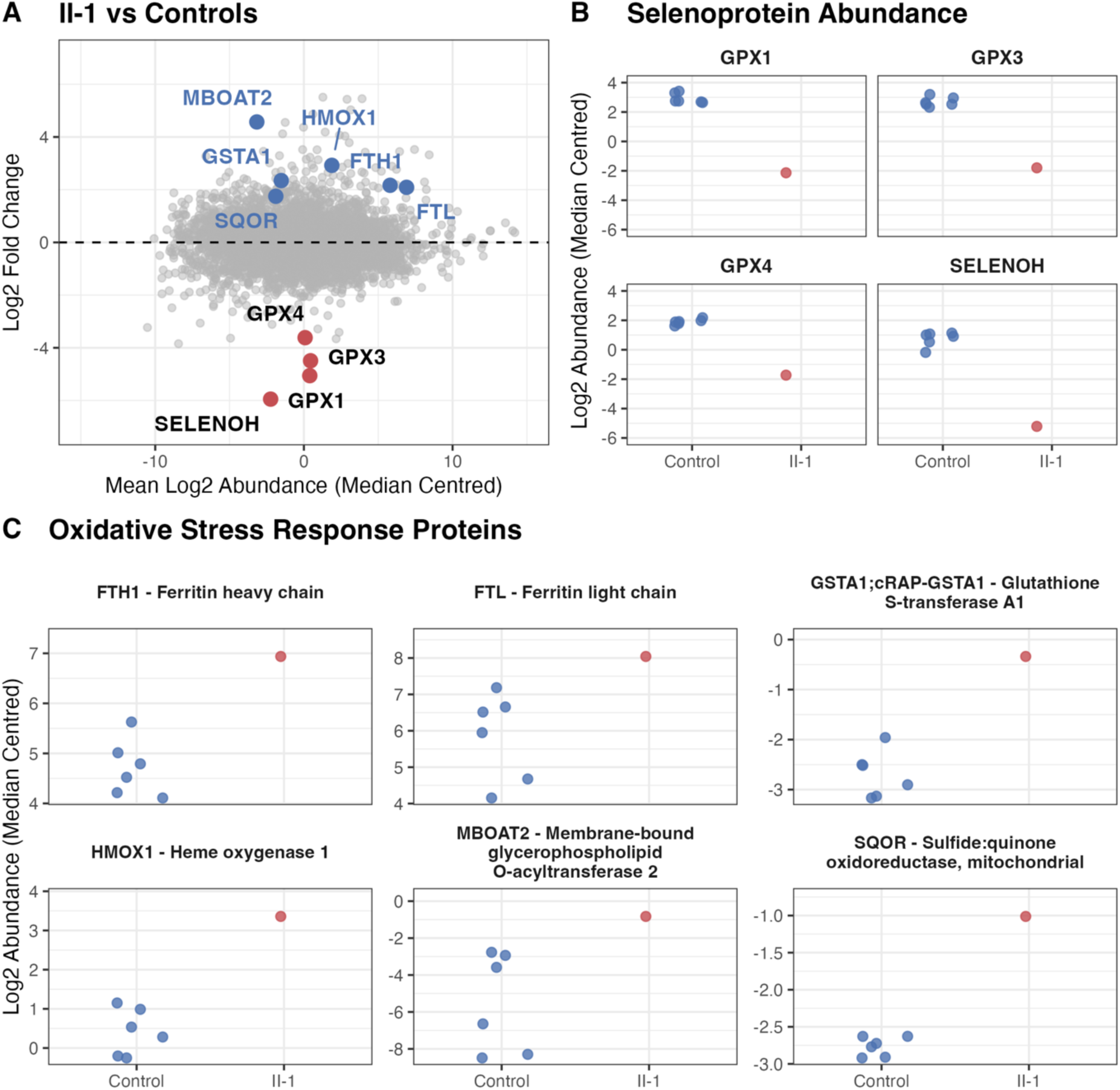
Cardiac Proteomics Analysis. Proteomics analysis of cardiac FFPE tissue from II-1 obtained at autopsy and six deceased female term babies without cardiac disease. **A:** MA plot depicting log2 normalised median-centred protein abundances in II-1 vs controls (red = downregulated selenoproteins, blue = upregulated oxidative response proteins and ferritin light and heavy chains). **B:** Key selenoproteins with large log2 fold changes in II-1 vs controls (GPX1 Log2FC -5.06, GPX3 Log2FC -4.49, GPX4 Log2FC -3.62, SELENOH Log2FC -5.95). **C:** Selected upregulated proteins involved in oxidative stress responses, iron sequestration or the inhibition of ferroptosis (FTH1, FTL, GSTA1, HMOX1, MBOAT2 and SQOR).

GPX4 is essential for the suppression of ferroptosis, an iron-dependent non-apoptotic cell death pathway involving lipid peroxidation and plasma membrane rupture^30–32^. The upregulation of ferritin (FTH1/FTL) is potentially indicative of increased iron sequestration as a compensatory mechanism to avoid ferroptosis in a failing heart with extreme GPX4 deficiency. MBOAT2 is a known inhibitor of ferroptosis and the upregulation of MBOAT2 may thus reflect a cellular response to protect against membrane peroxidation by altering the unsaturated fatty acid content of lipid membranes^32^. However, we acknowledge the limitation that MBOAT2 is transcriptionally regulated via the androgen receptor^32^ and that all control samples are from females. Additionally, SQOR is involved in the reduction of ubiquinone to generate ubiquinol, which acts as a lipid-peroxide scavenger to suppress ferroptosis^33^. Collectively these changes in protein abundance reveal a pronounced loss of key cardiac selenoproteins along with a compensatory response, perhaps to mitigate against ferroptosis due to GPX4 deficiency.

### Spectrum of Reported SECISBP2 Variants

We identified 18 biallelic variant pairs in patients with a *SECISBP2* deficiency disorder, representing a combination of homozygous and compound heterozygous genotypes (**Figure 7**)^19–22,34–38^. The location of *SECISBP2* variants is critical for determining their impact and the downstream phenotype. Some variants, such as p.Lys438Ter and p.Gln571Ter, are predicted to be null alleles that undergo NMD. Other variants have been shown to be functionally null. For instance, the p.Cys691Arg allele is expected to disrupt the folding of the L7Ae domain, abolishing protein function *in vitro* and causing a lethal phenotype in homozygous mouse models^22,29^. Upstream truncating variants (<Met300) are predicted to be hypomorphic due to transcript rescue from alternative translation initiation sites, thus preserving downstream functional domains. Crucially, we suggest that all previously reported patients carry at least one hypomorphic allele, resulting in partial protein function (**Figure 7, Supplementary Table 1**). In contrast, we predict that the homozygous c.1303-2A>G variant results in a near complete loss of functional SBP2 protein.

**Figure 7:**
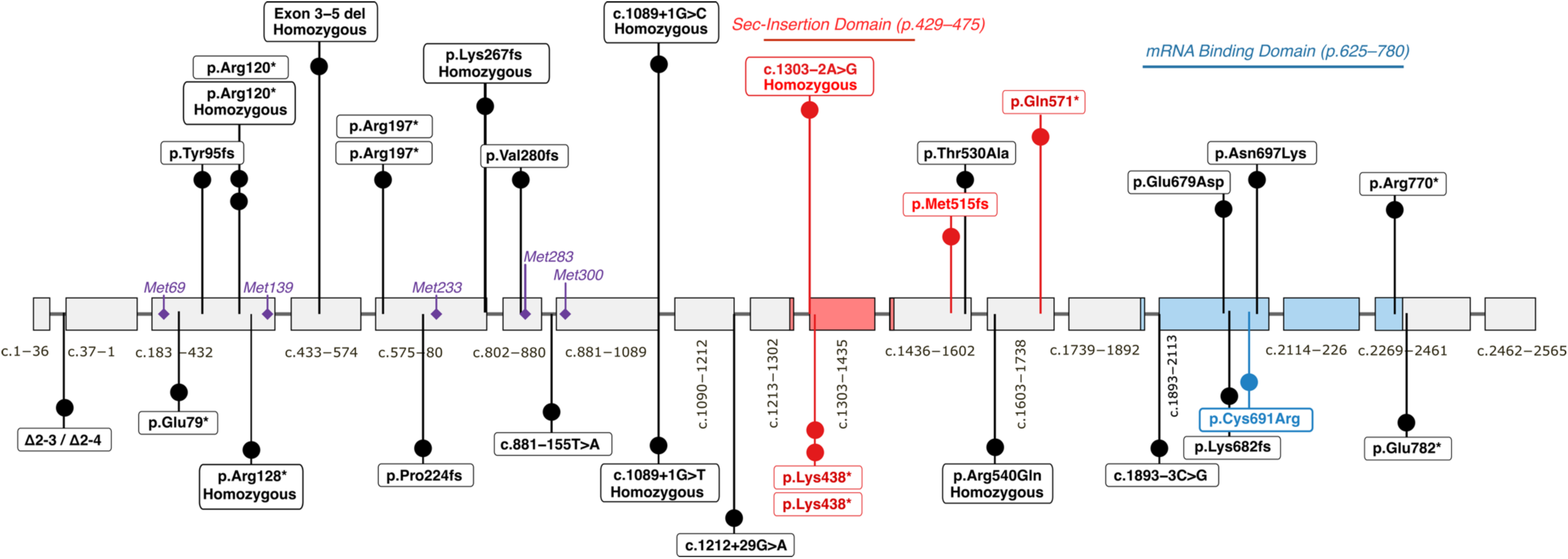
Pathogenic Biallelic *SECISBP2* Variants. Biallelic *SECISBP2* variants reported in the literature^19–22,34–38^. Variants in black are predicted to be hypomorphic, with some potential rescue to transcripts. Variants in red are predicted to undergo NMD. Possible alternative initiation translation sites are depicted in purple with methionines 1, 139, 233 and 300 representing the main alternative translation sites^36^. Exon 10 within the Sec-insertion domain (red), is located downstream of all alternative initiation sites. The p.Cys691Arg variant (blue) is functionally null as the resulting protein cannot bind SECIS mRNA *in vitro*^22,29^. All previous cases are predicted to carry at least one predicted hypomorphic allele.

## DISCUSSION

We report a homozygous canonical acceptor splice-site variant, present in four offspring with a severe lethal perinatal cardiomyopathy, expanding the phenotype spectrum of *SECISBP2* disease and representing the most severely affected family reported to date. These findings identify cardiomyopathy as a core feature of *SECISBP2* deficiency, providing the first *genetic* evidence that disruption of the selenoprotein synthesis pathway is sufficient to cause human heart failure.

*SECISBP2* deficiency causes pleiotropic phenotypes and the earliest reported cases had mild phenotypes. The first report described disrupted thyroid metabolism due to the impaired synthesis of the iodothyronine deiodinases in homozygous carriers of the p.Arg540Gln allele^19^. This variant impairs SBP2 binding to a specific subset of selenoprotein SECIS elements, with the disruption of DIO selenoprotein synthesis driving the reported thyroid phenotype^39^. In another study, the respective mouse mutant protein was found to be heat labile *in vitro* and to have reduced affinity for several SECIS elements, including *Dio1*^29^.

All previously reported cases of *SECISBP2* deficiency are compatible with life. Therefore, a fundamental question is understanding why the children in our family developed a lethal perinatal cardiomyopathy resulting in stillbirth or neonatal death. We suggest that rescue mechanisms allow for residual protein function in all previously reported cases, with affected individuals carrying at least one hypomorphic allele (**Figure 7, Supplementary Table 1**)^19–22,34–38^. The *SECISBP2* gene contains many alternative translation initiation sites, notably Met 139, 233 and 300, which generate different isoforms^21,22,34,40^. Further, the key functional domains are all situated downstream of these alternative translation initiation sites^15,36,40^. This may explain why patients with homozygous nonsense or truncating frameshift variants in upstream regions do not exhibit the fatal phenotype observed in homozygous knockout mouse models^20,21^.

Further, we suggest that the two homozygous splice variants reported by Stoupa et al. as null alleles (c.1089+1G>C and c.1089+1G>T) are likely hypomorphic^21^. While gross motor delays and muscle weakness were reported, there were no cardiac features, and ages at the last evaluation ranged from 22 months to 3.5 years. A close examination of the clinical data shows that the child with the homozygous c.1089+1G>C variant had serum GPX activity comparable to that of other reported patients with hypomorphic variants^21^. We propose that these two splice site variants result in out-of-frame exon 7 skipping, with partial transcript rescue via a cryptic donor at c.1009, generating a protein with a deletion of 27 amino acids from the 5’ of exon 7, p.Val337_Lys363del, upstream of the Sec-insertion domain. In support of this hypothesis, this cryptic donor site is observed in 0.4% of all GTEx and SRA samples. Other null alleles, such as p.Lys438Ter and p.Gln571Ter, have only been reported in compound heterozygous patients in combination with a less deleterious allele^19–21^.

Homozygous deletion of *Secisbp2* in mice results in embryonic lethality, with evidence of impaired development as early as embryonic day 8^18^. Further, other components of the selenoprotein system are essential for life. In mice, the homozygous deletion of the *Trsp* gene, which encodes the selenocysteine tRNA, results in embryonic lethality^41^ and the selective knockout of *Trsp* in cardiac and skeletal muscle causes acute myocardial failure^42^. Further, *GPX4* biallelic loss-of-function causes Sedaghatian-type spondylometaphyseal dysplasia (SSMD), a lethal autosomal recessive disorder characterised by cardiac conduction and central nervous system defects as well as mild limb shortening^43–46^. Aspects of SSMD bear striking parallels to the phenotype of the affected children in our family, with reports of heart block, cardiac structural defects and neonatal death with cardiorespiratory failure or cardiac arrhythmias in SSMD^43,44,46^. To highlight these parallels, one neonate with SSMD developed a sudden decrease in arterial oxygen saturation, bradycardia and ventricular tachycardia shortly after birth, dying the next day. Another affected sibling developed complete heart block and died shortly after birth from cardiorespiratory failure^46^.

This link with *GPX4* is important, as *SECISBP2* deficiency is known to adversely affect both *GPX4* mRNA stability and translation efficiency^17,29^. Consistently, we observed a very large decrease in cardiac GPX4 protein abundance in the first neonate that underwent autopsy (Log2FC -3.62) which we suggest is a likely contributor to the fatal cardiomyopathy. Given the pronounced loss of cardiac GPX4 protein abundance (>12-fold linear decrease) and the close phenotypic match with SSMD, we suggest that near total *SECISBP2* deficiency shares a similar pathology to SSMD. A mouse model carrying a patient-derived homozygous mutation in *SEPSECS*, another gene in the selenoprotein biosynthesis pathway, also exhibits a remarkably similar phenotype: the mice are born without obvious defects, but develop bradycardia and finally cardio-respiratory arrest several hours after birth^47^. In these mice, *Selenoh* and *Selenow* are downregulated, and genes like *Gsta4* are induced in the heart. Interestingly, overexpression of a selenium-independent GPX4 transgene rescued the otherwise selenoprotein-deficient mice, suggesting that lipid-peroxidation may be the underlying pathological pathway.

Previously reported cases of *SECISBP2* deficiency are associated with a predisposition to developing aortic aneurysms^20^. The proposed mechanism involves increased oxidative stress and aortic vascular smooth muscle cell death via ferroptosis due to the decreased synthesis of antioxidant selenoproteins^20^. In support of this theory, patient cells exhibited increased hydrogen peroxide production and membrane phospholipid peroxidation as well as decreased GPX1, GPX3, GPX4 and SELENOH protein abundance and decreased glutathione peroxidase and thioredoxin reductase activity^20^. A similar mechanism has also been proposed in the context of Keshan disease, a cardiomyopathy underpinned by dietary selenium deficiency^4^. Notably, Pei et al. observed decreased GPX1 and TXNRD1 protein abundance in cardiac tissue of patients with Keshan disease^48^. Consistently, we observed a pronounced loss of the selenoproteins SELENOH, GPX4, GPX3 and GPX1 in cardiac tissue from one homozygote.

GPX1 and GPX3 are involved in the reduction of hydrogen peroxide whereas GPX4 is a critical regulator of ferroptosis by reducing lipid hydroperoxides and protecting against lipid peroxidation^31,49^. The severe deficiency of these key selenoproteins supports an oxidative stress-mediated mechanism for cardiac pathology, possibly encompassing cardiac damage due to ferroptosis due to the loss of GPX4. We also observed an upregulation of FTH1, FTL, MBOAT2 and SQOR in patient cardiac tissue, which we interpret as a possible compensatory response to sequester iron and suppress ferroptosis in a state of GPX4 deficiency. MBOAT2 is notable as a recently identified GPX4-independent suppressor of ferroptosis via phospholipid remodelling^32^ and SQOR is involved in the suppression of ferroptosis via the hydrogen selenide-mediated reduction of ubiquinone to ubiquinol^33^. While it is difficult to fully distinguish compensatory changes due to heart failure versus ferroptosis-specific changes, we suggest that the loss of GPX4 is a major driver of the lethal cardiac phenotype arising due to *SECISBP2* deficiency in our family.

## CONCLUSION

The *SECISBP2* c.1303-2A>G variant causes a severe autosomal recessive selenoprotein deficiency. This is a fatal genetic condition that mirrors the embryonic lethality observed in *Secisbp2* knockout mice and shares close phenotypic and molecular parallels to SSMD, a genetic condition caused by the biallelic loss of GPX4. This is the first reported instance of a perinatally lethal cardiomyopathy due to biallelic variants in *SECISBP2*, illustrating, for the first time, that this gene is essential for human development.

## Data Availability

All data produced in the present study are available upon reasonable request to the authors and following appropriate ethics and legal approval.

## ACKNOWLEDGEMENTS

We sincerely thank the family who participated in this research for their significant contribution to advancing the scientific understanding of rare and inherited genetic diseases. This research was supported by the Medical Research Future Fund (MRFF) Cardiovascular Health Mission (# 2024269), a New South Wales Health Cardiovascular Capacity grant and philanthropic funding from Alexandra’s Mission and Heart of the Green in memory of Josh Avvenevole. JI, DGM and ERP are supported by National Health and Medical Research Council investigator grants (#2034308, #2009982, #2008376). JI and JM are supported by National Heart Foundation of Australia Future Leader Fellowships (#106732, #107192). This research was undertaken with the assistance of resources from the National Computational Infrastructure (NCI Australia), an NCRIS enabled capability supported by the Australian Government. We thank the Garvan Molecular Genetics (GMG) facility and the Garvan sequencing platform for their assistance with RNA extraction and sequencing for a subset of samples. Analysis was supported by the Centre for Population Genomics (Garvan Institute of Medical Research and Murdoch Children’s Research Institute) and was funded in part by an MRFF Genomics Health Futures Mission grant (#2008820).

The biobanking of patient samples by the Melbourne Children’s Heart Tissue Bank was supported by the Loti and Victor Smorgon Family Foundation and Royal Children’s Hospital Foundation.

## ETHICS DECLARATION

The Elusive Hearts research study was approved by the Royal Children’s Hospital Human Research Ethics Committee (97521). Clinical case report ethics approval was granted by the Sydney Children’s Hospitals Network Human Research Ethics Committee (HREC) (CCR2026/9).

