## Supplementary Table 1 for "*SECISBP2* Deficiency Causes a Lethal Perinatal Cardiomyopathy"

### SUPPLEMENTARY MATERIALS

#### Supplementary Table 1: Published *SECISBP2* Variants and the Corresponding Clinical Phenotype

| Genotype | Variant | VEP | Phenotype | Case Conclusion | Reference |
|---|---|---|---|---|---|
| Homozygous | c.1303-2A>G p.? | Splice acceptor | Lethal perinatal cardiomyopathy with intractable arrhythmias. | Complete biallelic loss of function (LoF). | This case. |
| Compound heterozygous | c.1312A>T p.(Lys438*) | Stop-gain | Sensorineural hearing loss, widening of the ascending thoracic aorta / aortic regurgitation and seizures. | Total LoF in one allele only, partial rescue of splice acceptor variant from cryptic acceptor site (activation of cryptic acceptor site 12bp downstream). | (Schoenmakers et al. 2023) |
| | c.1893-3C>G p.? | Splice acceptor | | | |
| Compound heterozygous | c.669del p.(Pro224Leufs*32) | Frameshift | Muscle weakness, fatigue, Raymund disease, developmental delay, raised T4, hearing problems, sun photosensitivity, testicular torsion, primary infertility, rotatory vertigo. | Partial rescue of LoF allele from alternate start codon<br>Production of hypomorphic protein from 2nd allele with a truncated protein. | (Schoenmakers et al. 2023, 2010) |
| | c.1090-155T>A p.? | Intronic | | | |
| Homozygous | c.382C>T p.(Arg128*) | Stop-gain | Abnormal thyroid hormone levels, aortic root dilatation, muscle weakness. | Partial rescue of homozygous LoF alleles from alternate start codon. | (Schoenmakers et al. 2023; Di Cosmo et al. 2009) |
| Homozygous | c.182+676_574+148del / c.(182+1_183-1)_(574+1_575-1)del | Deletion | Delayed growth, short stature, muscle weakness, incoordination, aortic dilatation, valvular insufficiency. | Partial rescue of homozygous LoF alleles from alternate start codon. | (Schoenmakers et al. 2023) |
| Compound heterozygous | c.1588A>G p.(Thr530Ala) | Missense | Patient 1: Abnormal thyroid hormone, delayed growth and bone maturation. Patient 2: Failure to thrive, abnormal thyroid hormones. | Total LoF one allele only, hypomorphic protein containing missense variant produced from 2nd allele. | (Stoupa et al. 2024) |
| | c.1711C>T p.(Gln571*) | Stop-gain | | | |
| Compound heterozygous | c.283del p.(Tyr95Ilefs*31) | Frameshift | Global developmental delay, failure to thrive, delayed bone maturation, autism, abnormal thyroid hormones, ADHD. | Partial rescue of both LoF alleles from alternate start codons. | (Stoupa et al. 2024) |
| | c.589C>T p.(Arg197*) | Stop-gain | | | |
| Compound heterozygous | c.838_839del p.(Val280Asnfs*3) | Frameshift | Hypotonia, autism, hearing deficit, non-verbal, abnormal thyroid hormones. | Partial rescue of LoF allele from alternate start codon transcript, production of hypomorphic protein from 2nd allele. | (Stoupa et al. 2024) |
| | c.2091C>A p.(Asn697Lys) | Missense | | | |
| Homozygous | c.1089+1G>C p.? | Splice donor | Failure to thrive, global developmental delay, hypotonia, short stature. | Affects splicing - out-of-frame exon 7 skipping, with partial rescue via activation of a cryptic donor at c.1009, resulting in a protein product containing a deletion of | (Stoupa et al. 2024) |

| | | | | | |
|---|---|---|---|---|---|
| | | | | 27 amino acids from the 5' of exon 7, p.(Val337_Lys363del). | |
| Homozygous | c.1089+1G>T p.? | Splice donor | Abnormal thyroid hormones, developmental delay, muscle weakness, moderate sensorineural hearing loss. | Affects splicing - out-of-frame exon 7 skipping, with partial rescue via activation of a cryptic donor at c.1009, resulting in a protein product containing a deletion of 27 amino acids from the 5' of exon 7, p.(Val337_Lys363del). | (Stoupa et al. 2024) |
| Homozygous | c.358C>T p.(Arg120*) | Stop-gain | Hypotonia, abnormal thyroid hormones, ADHD, learning difficulties, language delay. Unrelated sickle cell disease. | Partial rescue of homozygous LoF alleles from alternate start codon transcript. | (Stoupa et al. 2024) |
| Homozygous | c.1619G>A p.(Arg540Gln) | Missense | Abnormal thyroid hormones. | Production of hypomorphic protein containing missense variant from homozygous alleles. | (Dumitrescu et al. 2005) |
| Compound heterozygous | p.Lys438* | Stop-gain | Abnormal thyroid hormones and growth delay. | LoF in one allele (p.Lys438*).<br>Partial LoF in the splice donor allele, 52% of the transcripts from the splice donor allele are reported to be abnormally spliced (Dumitrescu et al. 2005). | (Dumitrescu et al. 2005) |
| | c.1212+29G>A | Splice donor | | | |
| Compound heterozygous | c.358C>T (p.Arg120*) | Stop-gain | Abnormal thyroid hormones, delayed bone maturation, congenital myopathy, impaired motor function. | Partial rescue of Arg120* allele from alternate start codons.<br><br>The p.Arg770* variant may avoid NMD due to an in-frame rescue transcript: the naturally occurring ex15-16Δ transcript will not include the variant and encoded a PTC, resulting in the production of a truncated protein. | (Azevedo et al., 2010) |
| | c.2308C>T (p.Arg770*) | Stop-gain | | | |
| Homozygous | c.800_801insA p.(Lys267Lysfs*2) | Frameshift | Abnormal thyroid hormones, ADHD and muscle weakness. | Partial rescue from alternate start codons. Indeed, a shorter SBP2 isoform was observed on western blots (Çatli et al. 2018). | (Çatli et al. 2018) |
| Compound heterozygous | c.235C>T p.(Glu79*) | Stop-gain | Abnormal thyroid hormones, short stature and delayed bone maturation. Mild conductive hearing loss. | Partial rescue of Glu79* allele from alternate start codons as suggested by the authors (Hamajima et al. 2012). | (Hamajima et al. 2012) |
| | c.1529_1541dupCCAGCGCCCCACT p.(Met515Glufs*48) | Frameshift | | | |

| | | | | | |
|---|---|---|---|---|---|
| Compound heterozygous | c.589C>T p.(Arg197*) | Stop-gain | Abnormal thyroid hormones (high T4, low-normal T3, high reverse T3, normal TSH). Growth and developmental delay. Decreased serum glutathione peroxidase enzymatic activity. | Partial rescue of the p.(Arg197*) allele. Two shorter isoforms generated from Met 233 and Met 300 were observed *in vitro* (Fu et al. 2020). | (Fu et al. 2020) |
| | c.2037G>T p.(Glu679Asp) | Missense | | | |
| Compound heterozygous | c.2045_2048delAACA p.(Lys682Thrfs*2) | Frameshift | Abnormal thyroid hormones (high T4, low T3, high reverse T3, slightly elevated TSH). Growth and developmental delay. Decreased serum glutathione peroxidase enzymatic activity. | For both alleles, isoforms with a truncated C-terminus were observed *in vitro* suggesting that both variants escape NMD (Fu et al. 2020).<br><br>In-frame truncated rescue transcripts exist for both alleles. For the p.(Glu782*) variant the ex15-16Δ transcript will not include the variant and encoded a PTC. For the p.(Lys682Thrfs*2) variant, the cryptic chr9:8935071-89357410 splice junction and the ex13-14Δ transcripts will not include the variant and encoded a PTC. | (Fu et al. 2020) |
| | c.2344C>T p.(Glu782*) | Stop-gain | | | |
| Compound heterozygous | c.2071T>C p.(Cys691Arg) | Missense | Global developmental delay, short stature, high T4, low T3, normal TSH. Muscle weakness, mild bilateral high-frequency hearing loss. | The allele that results in exon 2-4 skipping is hypomorphic, with rescue from alternative start codons. The p.(Cys691Arg) variant was not observed to yield a full length protein, and was suggested to result in proteasomal degradation (Schoenmakers et al. 2010). Based on mouse models, the p.(Cys691Arg) variant is proposed to be a functionally null allele (Zhao et al. 2019). | (Schoenmakers et al. 2010) |
| | Transcript described as lacking exons 2-4 or 3-4 | Splice variant | | | |
